# Bladder-resident bacteria associated with increased risk of recurrence after electrofulguration in women with antibiotic-recalcitrant urinary tract infection

**DOI:** 10.1101/2024.07.03.24309902

**Authors:** Jashkaran G. Gadhvi, Parker R.M. Kenee, Kevin C. Lutz, Fatima Khan, Qiwei Li, Philippe E. Zimmern, Nicole J. De Nisco

## Abstract

**Background:** Antibiotic-recalcitrant recurrent urinary tract infection (rUTI) is has become increasingly observed in postmenopausal women. Therefore, when standard antibiotic therapies have failed, some elect electrofulguration (EF) of areas of chronic cystitis when detected on office cystoscopy. EF is thought to remove tissue-resident bacteria that have been previously detected in the bladder walls of postmenopausal women with rUTI. We hypothesized that increased bladder bacterial burden may be associated with incomplete rUTI resolution following EF.

**Methods:** Following IRB approval, bladder biopsies were obtained from 34 consenting menopausal women electing EF for the advanced management of rUTI. 16S rRNA FISH was performed using both universal and *Escherichia* probes and tissue-resident bacterial load was quantified. Time to UTI relapse after EF was recorded during a six-month follow-up period and the association of bladder bacterial burden and clinical covariates with UTI relapse was assessed.

**Results:** We observed bladder-resident *Escherichia* in 52% of all participants and in 92% of participants with recent *E. coli* UTI. Time-to-relapse analysis revealed that women with high bladder bacterial burden as detected by the universal probe had a significantly (*p*=0.035) higher risk of UTI within six months of EF (HR=3.15, 95% CI: 1.09-9.11). Interestingly, bladder-resident *Escherichia* was not significantly associated (*p*=0.26) with a higher risk of UTI relapse (HR= 2.14, 95% CI: 0.58-7.90).

**Conclusions:** We observed that total bladder bacterial burden was associated with a 3.1x increased risk of rUTI relapse within six months. Continued analysis of the relationship between bladder bacterial burden and rUTI outcomes may provide insight into the management of these challenging patients.

## Introduction

Urinary tract infections (UTI) are among the most common bacterial infections and can progress into more serious conditions like recurrent UTI (rUTI) and pyelonephritis (1, 2). rUTI, defined as two UTI episodes in 6 months or three UTI episodes in 12 months, has become a significant clinical challenge in the older female population (3, 4). Ikäheimo *et al.* followed 179 women for 12 months after initial presentation to a clinic for treatment of UTI to assess the incidence of rUTI, and within that period 53% of individuals ≥ 55 y/o experienced rUTI (5). A separate study found a high incidence of resistance or allergy to multiple front-line UTI antibiotics in postmenopausal women with rUTI (6). In older women, several factors are associated with a higher risk of rUTI, including institutionalization, catheterization, incontinence, antimicrobial exposure, and decreased functional status (3). Of these factors, the contribution of prior antimicrobial exposure to rUTI is particularly concerning, as the development of antibiotic resistance reduces the number of options available to clinicians for treating rUTI and leaves patients vulnerable to pyelonephritis and urosepsis (2, 7, 8). Therefore, a deeper understanding of how bacterial pathogens colonize and persist in the urinary tract and the role of bladder-resident bacteria in the rUTI patient outcomes is needed.

Uropathogenic *Escherichia coli* (UPEC) is the most common causative agent of UTI. Studies in mice have shown that UPEC forms intracellular bacterial communities (IBCs) and quiescent intracellular reservoirs (QIR) that can seed future infections (9-12). Additional studies in mice and in vitro systems have also found that uropathogens *Klebsiella pneumoniae* and *Enterococcus faecalis* can also invade bladder epithelial cells (13-15). The presence of tissue-embedded bacteria in the bladders of human patients with rUTI was confirmed in our study of 14 bladder biopsies from postmenopausal women with rUTI. Fluorescence *in situ* hybridization (FISH) using universal bacterial 16S rRNA demonstrated that bladder-resident bacteria were present in 11 out of 14 patients in all layers of the urothelium and even within the sub-urothelial mesenchymal regions (16). One treatment modality for women with antibiotic-recalcitrant rUTI that exploits our understanding of uropathogen tissue invasion is cystoscopy with electrofulguration (EF) directly to areas of visible chronic cystitis (17, 18). Retrospective assessment of 95 women undergoing EF for rUTI from 2004 to 2016 indicated 65% of women experienced a reduction in their number of UTIs per year when followed for an average of 4.9 years EF (17).

However, EF procedures do not resolve rUTI in some patients, thus prompting the question of whether the burden of bladder-resident bacteria may influence the early EF outcomes. Furthermore, the identity of these bladder-resident bacteria remains unknown as previous work employing FISH to identify bladder-resident bacteria has only utilized universal, and not genus-specific, bacterial probes (16, 19). To address these questions, systematic analysis was performed on bladder biopsies taken from 34 post-menopausal women prior to undergoing EF for antibiotic-recalcitrant rUTI. Tissue samples were analyzed with FISH probes to both universal 16s rRNA sequences, and to *Escherichia spp.* specific sequences. The biopsies were independently analyzed to quantify and characterize genus-specific detection of bacteria within the bladder wall. This study represents the first successful genus-specific typing of bladder-wall resident *Escherichia* in postmenopausal women suffering from uncomplicated antibiotic-recalcitrant rUTI. Strikingly, we found that elevated bladder tissue bacterial burden correlated with an increased risk of rUTI relapse within the first 6 months after EF. Therefore, this observation identifies a new prognostic bladder indicator, easily obtained through biopsy during the EF procedure, that could influence the early management following EF for improved clinical outcomes.

## Methods

### Patient Recruitment

Following institutional review board (STU-042018-072, MR 17-120) approval, eligible postmenopausal women undergoing EF were recruited from the Urology clinic at the University of Texas Southwestern Medical Center. EF was indicated in menopausal women with a history of uncomplicated antibiotic-recalcitrant rUTI, completely negative upper and lower urinary tract evaluations, and areas of visible chronic cystitis on office flexible cystoscopy. Exclusion criteria included neurogenic bladder, pelvic procedure for incontinence or pelvic organ prolapse within 6 months prior, uncontrolled diabetes, >stage 2 prolapse, voiding dysfunction, ongoing chemotherapy, renal insufficiency, use of indwelling catheters, intermittent catheterization, no cystitis found on cystoscopy, or any upper urinary tract anomaly explaining rUTI.

### Cystitis Stage Classification

Cystitis stages were devised for in-house classification of chronic cystitis bladder involvement. Based on cystoscopy findings, areas of chronic cystitis can be categorized according to their extension to the trigone (stage 1 or trigonitis), trigone and bladder base (stage 2), trigone, bladder base and lateral walls of the bladder (stage 3) or the rest of the bladder (stage 4 or pancystitis) (20).

### Sample Collection and Preservation

Bladder biopsies were obtained from menopausal women who met the study criteria and provided informed consent. For Fluorescence in Situ Hybridization (FISH) analysis, the collected biopsies were immediately placed in 4% paraformaldehyde and fixed for 6 hours at room temperature or overnight at 4°C. The fixed biopsies were then paraffin embedded and longitudinally sectioned (5 µm) using sterile solutions and equipment as described previously (21).

### Fluorescence in situ hybridization (FISH)

All staining steps were performed in aseptic conditions as previously reported (21, 22). Sequential sections were incubated overnight at 55°C with either: 10nM 16S rRNA-647 (Universal) + 10nM *Esco473*-488 (*Escherichia*), 10nM *Esco473*-488 (*Escherichia*) alone or 10nM scramble-647 + 10nM scramble-488 probe sets diluted in hybridization buffer (23). After washing, sections were counterstained with 1 μg/mL Hoechst, washed and then mounted. Confocal microscopy was performed on a Zeiss LSM800 with a 63x objective.

### Image Analysis

Images were processed and analyzed with ImageJ. Ten random images including the urothelial region, the lumen and the basal region were taken for each biopsy tissue. Anatomical regions (i.e. lumen, urothelium, lamina propria) present with the images were defined were defined using Hematoxylin and Eosin (H&E) staining on serial sections. Dotted lines represent the delineation between the urothelium and the sub-urothelium. After imaging was performed, two separate, clinically blinded or unaware of the EF outcomes evaluators performed analysis and quantification of bacteria present within each tissue layer. Quantified bacterial counts were then used for statistical analyses of bladder bacterial burden.

### Statistical Analysis

Sample sizes balanced feasibility of biopsy acquisition with statistical power for detecting appropriate effect sizes. For example, for the survival analysis a sample size of N=31 provides 80% power to detect large effect sizes (HR>2) (24). All statistical analyses were performed in GraphPad Prism 8.1.0 or in R Studio version 4.0.2 with a level of significance of a 0.05. Pairwise associations between continuous variables were measured using Spearman’s correlation coefficient with *p*-values generated by permutation. Differences between continuous variables by group were analyzed Mann-Whitney-Wilcoxon test (for two group comparison) or Kruskal–Wallis test (for multiple group comparison).

### Time-to-Relapse Analysis

UTI episodes were recorded in a six-month follow-up study in which patients were followed through chart review in electronic medical records and phone calls. UTI relapse was defined as one symptomatic, culture-proven UTI episode within six months after the EF procedure. Survival analysis was performed on the relapse group using Kaplan-Meier analysis and Cox proportional hazards models. All model assumptions for Cox regression including proportional hazards, influential observations, and linearity (for quantitative predictors) were carefully evaluated using the Schoenfeld test, deviance residual plots, and Martingale residual plots, respectively. Participants were dichotomized about the median for Universal bacteria and by presence or absence for the *Escherichia* probe. No other variables in our data set were associated with both the independent and dependent variables, Universal bacteria and rUTI relapse, respectively. This implies that there are likely to be no confounders between Universal bacteria and rUTI relapse in our data set. The Mantel-Cox log-rank test was used to compare survival curves in the Kaplan-Meier analysis to determine if groups differed with respect to time-to-relapse. We then performed Cox regression to account for clinical covariates. Because some variables were correlated and the number of events per covariate was about two, we performed regularization on the Cox regression model with all the covariates using LASSO from the glmnet library in R Studio. Penalization techniques such as LASSO are recommended especially when the number of events per covariate is less than four or five (25). To identify confounders, we determined if any variables were associated with both the independent and dependent variables: universal bacteria and rUTI relapse, respectively. We used the p-value and 95% confidence interval of the hazard ratio corresponding to each covariate to quantify their association with the hazard and survival time.

### Ethics Statement

This study was performed with approval from the Institutional Review Boards of the University of Texas Southwestern Medical Center and the University of Texas at Dallas (STU-042018-072, MR 17-120). Informed, written consent was obtained from all patients before performance of study procedures or collection of study-related data.

## Results

### Patient characteristics and urine culture history

All patients were postmenopausal except one who was perimenopausal (PNK036) with a mean age of 69 years and age range from 44-92 years (**Table 1**). Their demographic characteristics are presented in Table 1. The participants had body mass indices ranging from 19 – 47.7 (median, 25.8). Ten women (29%) suffered from Type 2 diabetes mellitus and the median parity of the cohort was two (**Table 1**). Most of the women (76%) had gynecologic surgical history ranging including Cesarean section, abdominal hysterectomy, vaginal hysterectomy, sling placement, and nephrectomy. Of the 34 patients, three had previously undergone at least one EF procedure (PNK 025, PNK027, and PNK033). Twenty-three participants (68%) had allergies to antibiotics. Specifically, recorded allergies were to Nitrofurantoin (24%), Fluoroquinolones (29%), Macrolides (13%), Sulfonamides (41%), Aminoglycosides (3%), Penicillin (15%) and Cephalosporin (9%). Fifteen participants (44%) had allergies to multiple antibiotics (**Table 1**).

**Table 1.** Patient clinical characteristics. Overview of cohort clinical characteristics including age, body mass index (BMI), diabetes, parity, gynecologic surgical history, and antibiotic allergies. SD: Standard deviation; IQR: Interquartile range. *Patients with multiple factors in a single category were counted separately for summation of each factor.

| **Clinical Characteristic** |  |
| --- | --- |
| **Mean age, years (SD)** | 69 (10.9) |
| Range | 44 - 92 |
| **Median BMI, kg/m<sup>2</sup> (IQR)** | 25.8 (23.1 – 30.8) |
| Obese (BMI ≥ 30) | 9 (26%) |
| **Diabetic (all type II)** | 10 (29%) |
| **Parity** | 31 (91%) |
| Median (IQR) | 2 (1 – 3) |
| **Gynecologic Surgical History\*** |  |
| Any Surgery | 26 (76%) |
| Cesarean Section | 8 (24%) |
| Abdominal Hysterectomy | 8 (24%) |
| Vaginal Hysterectomy | 6 (13%) |
| Sling Placement | 9 (26%) |
| Nephrectomy | 2 (6%) |
| Other (BSO, cancer resection, Hysteroscopy, Cystectomy) | 18 (53%) |
| **Prior EF procedure** | 3 (9%) |
| **Antibiotic Allergies\*** |  |
| Any | 23 (68%) |
| Multiple | 15 (44%) |
| Nitrofurantoin | 8 (24%) |
| Fluoroquinolones (Ciprofloxacin, Levofloxacin) | 10 (29%) |
| Macrolides (Azithromycin, Erythromycin, Clindamycin) | 6 (13%) |
| Sulfonamides | 14 (41%) |
| Aminoglycosides (Gentamicin, Tobramycin) | 1 (3%) |
| Penicillin | 5 (15%) |
| Cephalosporin | 3 (9%) |

**Table 2** provides a description of the stage classifications of chronic cystitis used to stratify patients. Eight women were classified as Stage 1 (24%), nine as Stage 2 (26%), two as Stage 3 (6%), and nine as Stage 4 (44%). We also evaluated available urine culture history within one year prior to EF and found that 21 women (62%) had a history of *E. coli* UTI, 10 (30%) had *Klebsiella pneumoniae* UTI, and 8 (24%) had *E. faecalis* UTI (**Table 2**). All participants had taken at least one course of antibiotic therapy within one year prior to the study with many having had several courses without durable resolution of their rUTI condition. Of the 34 patients, 16 had a UTI episode requiring antibiotic treatment within 6 months post-EF.

**Table 2.** Cystitis stages, patient urine culture and antibiotic history within one year prior to EF. ESBL: extended spectrum beta-lactamase. NIT: nitrofurantoin, CIP: Ciprofloxacin, AMC: Amoxicillin-Clavulanate, FOF: Fosfomycin, DOX: Doxycycline, GEN: Gentamicin, LVX: Levofloxacin, CRO: Ceftriaxone, TMP/SMX: trimethoprim/sulfamethoxazole. Recurrent UTI relapse data for patients experience UTI within 6 months of electrofulgurations (EF) was denoted by (+) and not as (-).

|  | **Cystitis Stage** | **Urine Culture History (≤1 yr. prior to EF)** | **Antibiotic History (≤1 yr. to EF)** | **Relapse ≤6 mo.** |
| --- | --- | --- | --- | --- |
| PNK016 | 4 | *P. aeruginosa*, *P. mirabilis*, *M. morganii*, *E. faecalis* | Cefepime (IV), FOF, Amikacin, Cephalexin, Ertapenem, GEN | + |
| PNK017 | 4 | *E. coli*, *Enterobacter spp.* | CIP, Cephalexin, NIT, TMP/SMX | + |
| PNK018 | 1 | *E. coli*, *E. faecalis*, *K. oxytoca* | CIP, Ertapenem (IV), FOF, Cefuroxime, Cefdinir, NIT, AMC | + |
| PNK019 | 4 | *E. coli*, *K. pneumoniae*, *E. cloacae* | FOF, Cephalexin, AMC | - |
| PNK020 | 3 | *E. coli* | NIT, Cephalexin | - |
| PNK021 | 4 | *K. pneumoniae*, *E. coli* | CRO, Amoxicillin, Cephalexin, TMP/SMX, Cefazolin | + |
| PNK022 | 2 | *E. coli* | FOF, Ertapenem, Azithromycin, CIP, NIT, Meropenem | - |
| PNK023 | 3 | *K. pneumoniae* | Cefoxitin, DOX, Cefuroxime, FOF | + |
| PNK024 | 4 | *S. epidermidis*, *K. pneumoniae* | LVX, CIP, GEN, NIT | - |
| PNK025 | 4 | *E. coli*, *K. pneumoniae* | FOF, Cefazolin, TMP/SMX, NIT | + |
| PNK026 | 1 | *ESBL E. coli* | FOF, Meropenem (IV), GEN (IV), Cephalexin, Rifaximin | + |
| PNK027 | 4 | *K. pneumoniae*, *E. coli* | Levaquin, CIP, TMP/SMX, AMC, Amoxicillin, Cefazolin | - |
| PNK028 | 4 | *E. coli* | Cephalexin, CRO | - |
| PNK029 | 1 | *E. faecalis* | Levaquin | - |
| PNK031 | 2 | *E. coli*, *K. pneumoniae*, *K. aerogenes* | CIP, NIT, TMP/SMX, DOX, CRO | - |
| PNK032 | 1 | *E. coli* | NIT, Cefuroxime | - |
| PNK033 | 2 | *E. faecalis* | CIP, Levaquin, Amoxicillin, FOF, Cephalexin, TMP/SMX | + |
| PNK034 | 2 | *E. faecalis*, *K. pneumoniae* | Cephalexin, NIT | + |
| PNK035 | 4 | *E. coli* | AMC, NIT, Amoxicillin, TMP/SMX, Cefazolin | - |
| PNK036 | 2 | *S. agalactiae*, *K. aerogenes*, *K. pneumoniae* | AMC | - |
| PNK039 | 2 | none | GEN | - |
| PNK040 | 2 | *E. coli* | Aztreonam, NIT | - |
| PNK041 | 4 | *E. coli* | NIT, TMP/SMX | + |
| PNK042 | 4 | none | NIT | - |
| PNK043 | 4 | *E. coli* | Cephalexin, CIP, Cefuroxime | + |
| PNK044 | 4 | *K. pneumoniae*, *E. coli* | NIT, Cephalexin, CIP, TMP/SMX | + |
| PNK045 | 2 | *E. faecalis*, *E. coli* | LVX, NIT | - |
| PNK046 | 1 | *S. agalactiae* | NIT, CIP | + |
| PNK047 | 2 | none | NIT, Cephalexin | - |
| PNK048 | 1 | none | Cephalexin, CIP, NIT | - |
| PNK049 | 4 | *K. pneumoniae*, *E. coli*, *Citrobacter braaki*, *E. faecalis* | Amoxicillin, CIP | + |
| PNK050 | 1 | *E. faecalis*, *S. epidermidis* | TMP/SMX | + |
| PNK051 | 1 | *E. coli* | NIT | - |
| PNK052 | 4 | *E. coli* | LVX, TMP/SMX, CIP, Cefuroxime | + |

### Genus specific FISH reveals *Escherichia coli* bacterial communities within the bladder wall

Using universal probes for 16S rRNA gene, we detected bacteria within the bladder wall of 31/34 patients. Of these 34 patients, the universal FISH quantification had been reported for 11 patients in a previous study published by our group (26). **Figure 1A** shows representative micrographs from various stages (1, 2 and 4) of cystitis. Bacteria were present not just in the bladder urothelium, but also in the lumen and the sub-urothelial regions. Because *E. coli* is the most common causative agent of UTI and rUTI, we used a previously validated 16S rRNA probe for *Escherichia* (*Esco473*) to identify potential tissue-resident *E. coli* in a subset of the biopsies (23). We first performed our own in-house validation and confirmed that the *Esco473* probe was able to label *E. coli* isolated from patient PNK007 and not *Staphylococcus epidermidis* (**Figure S1**). Then we tested if the *Esco473* probe could detect potential *E. coli* within our bladder biopsies (**Figure 1B**). We found *Escherichia spp*. present in 12/23 (52%) of the biopsies and in 12/13 biopsy sections from women with recent history of *E. coli* UTI. We further confirmed the colocalization of both the universal as well as the *Escherichia* probe within biopsy tissues – PNK041 (**Figure 2A)**. We also observed instances, especially in women without recent history of *E. coli* UTI, where only the universal probe was detected suggesting that these bladder-resident bacteria do not belong to the genus *Escherichia* and may represent other genera of uropathogenic bacteria or potentially members of the urinary microbiota – PNK046 (**Figure 2B).**

**Figure 1.**
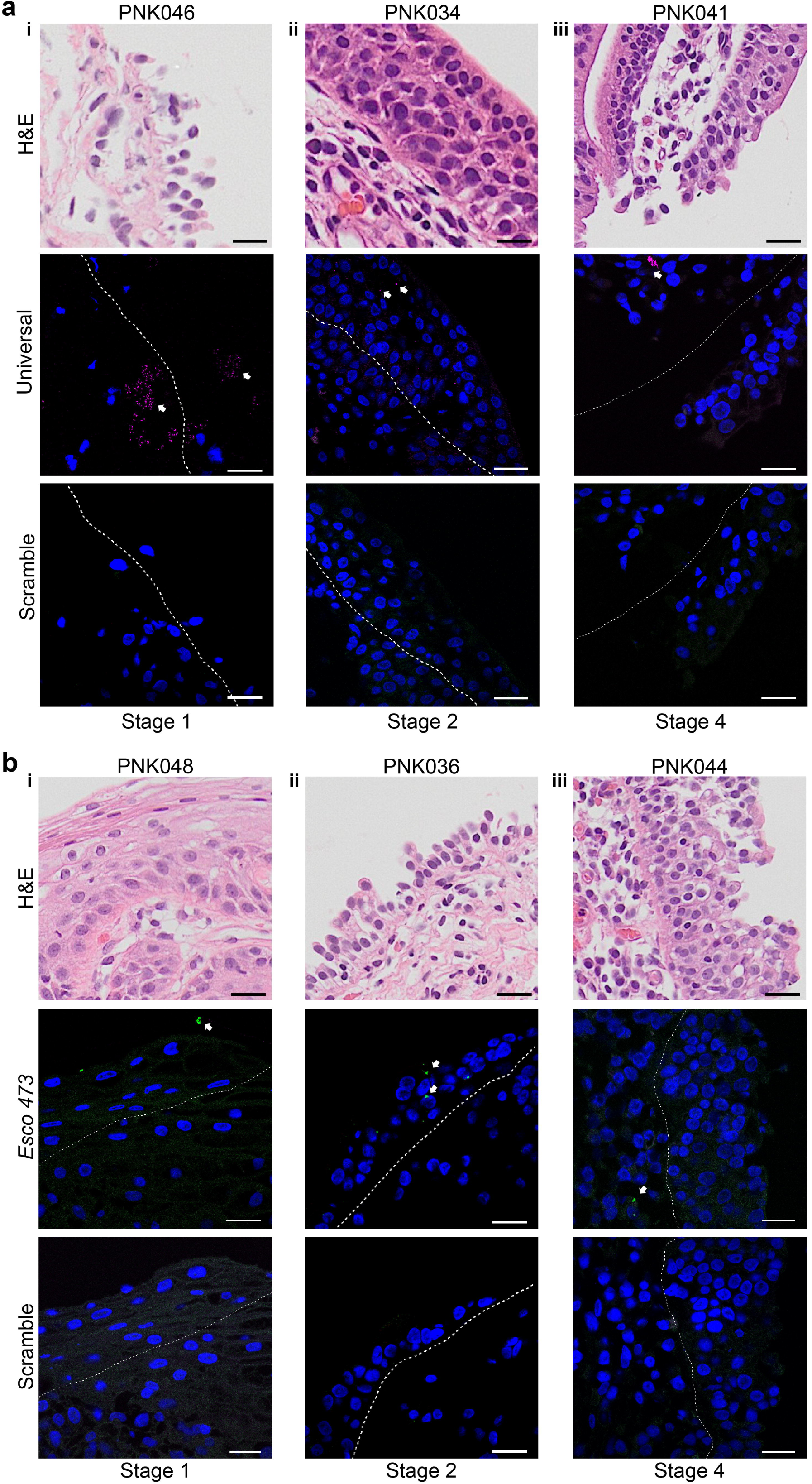
Genus specific FISH reveals presence of *Escherichia* in bladder tissues independent of cystitis stage. Representative micrographs show Universal probe (**A**) in magenta and *Escherichia* specific probe (**B**) in green detects tissue-resident bladder bacteria in all stages of cystitis. Scale bar: 20μm.

**Figure 2.**
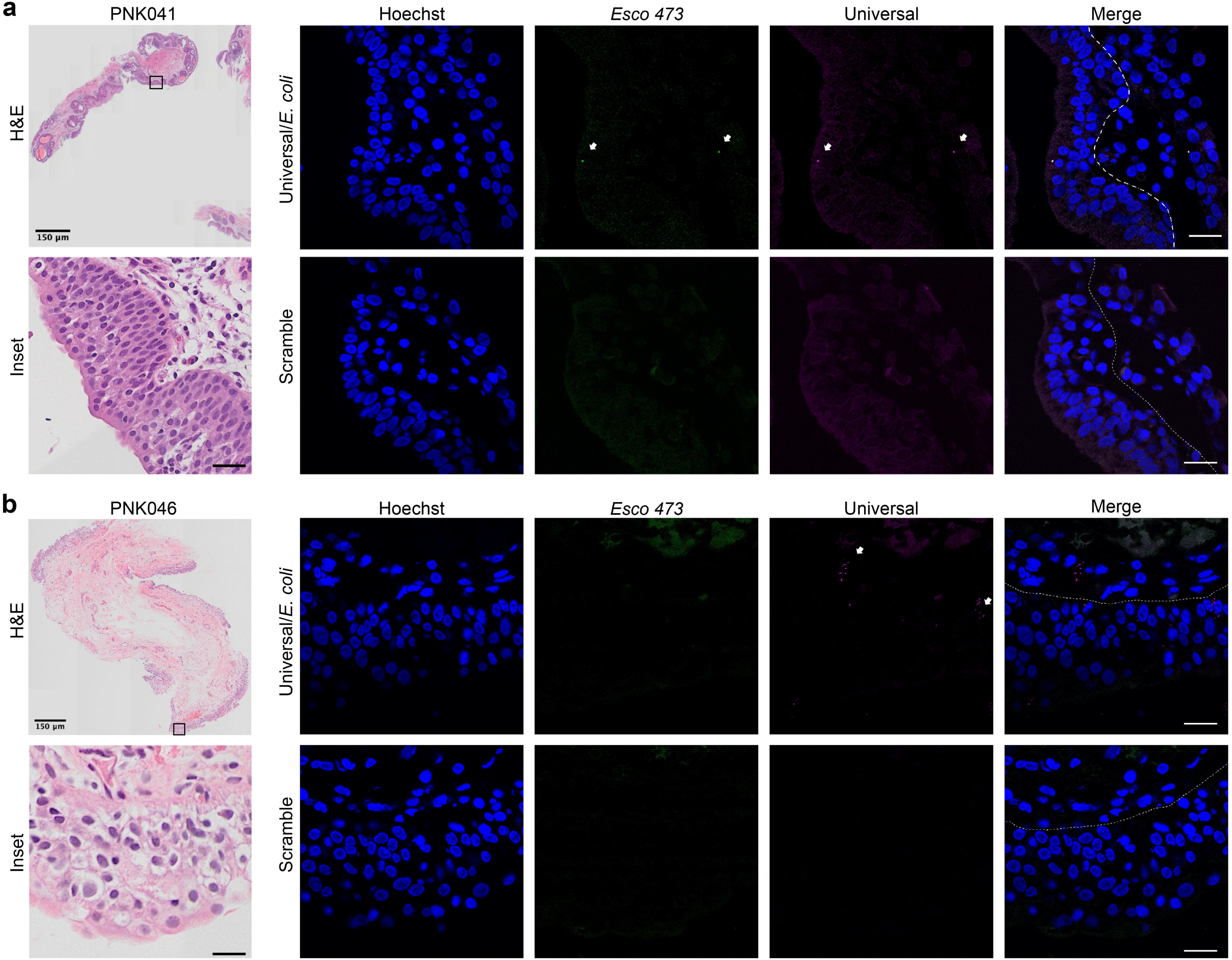
Co-hybridization with *Escherichia* and Universal probes allows simultaneous detection. (**A**) Bacteria can be detected using both the *Escherichia* and the Universal probe simultaneously on the bladder biopsy sections. (**B**) Co-hybridization demonstrates that the tissue resident bacteria in PNK046 are not *Escherichia.* Scale bar: 20μm.

### Bacterial burden and localization within the bladder wall

We then quantified the bladder resident bacteria within each layer of the urothelium to better understand the bladder burden of bacteria detected by the universal and *Esco 473* probes and the tissue distribution. The median total bladder bacterial burden, inclusive of lumen, urothelium, and suburothelium, detected by the Universal probe was 19 (IQR: 3-38.75) while the median total number of *Escherichia* was two (IQR: 0-18) (**Figure 3**). The largest bacterial burden detected by the Universal probe was in PNK046 with 368 bacteria, whereas the largest *Escherichia* community detected was in PNK041 with 26 bacteria (**Figure 3**). Interestingly, no *Escherichia* populations were observed in PNK046. However, this patient had no recent history of *E. coli* UTI, and their most recent UTI was suspected to be caused by *Streptococcus agalactiae.* (**Table 2**).

**Figure 3.**
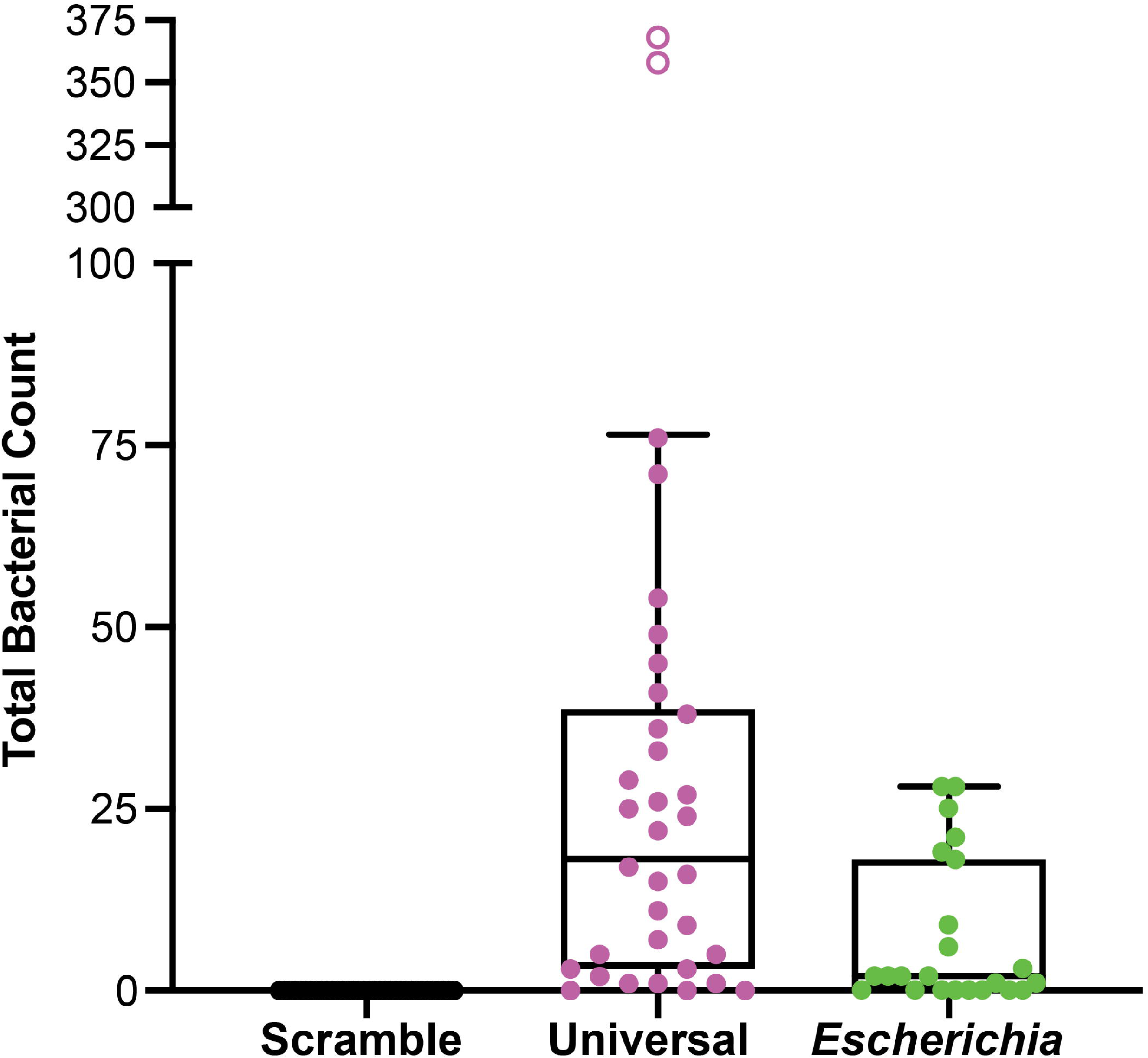
Quantification of total Universal and *Escherichia* bacterial load. Total bacterial count detected per patient biopsy with the Universal (magenta) and *Escherichia* (green) probes. Whiskers extend to the furthest data that are within 1.5 times the IQR. Outliers lie outside 1.5 times the IQR and are indicated by solid black dots outside of the whiskers when present. Each box indicates the IQR. The median is indicated by the solid line inside each box..

To ascertain the location of these communities of tissue-associated bacteria, we quantitated the bacteria present in lumen, urothelium, and suburothelium in PNK027-PNK052. Bacterial community tissue localization for PNK016-PNK026 was reported previously in Ebrahimzadeh et al (26). Bacterial communities were observed at all layers of the urothelial mucosa, from the bladder lumen to the urothelial and sub-urothelial layers (**Figure 4**). Tissue-resident (i.e., urothelial, or sub-urothelial) bacteria were detected by the universal probe in 87% of patients and their community sizes ranged from a single bacterium to large communities of up to 45 bacteria (**Figure 4A**). The largest urothelial and sub-urothelial communities detected by the Universal probe were observed in PNK046 (N=181) and PNK050 (N=21), respectively (**Figure 4A**). Luminal bacteria were detected by the universal probe in 43% of patient samples with PNK046 having the largest luminal bacterial community (N=149).

**Figure 4.**
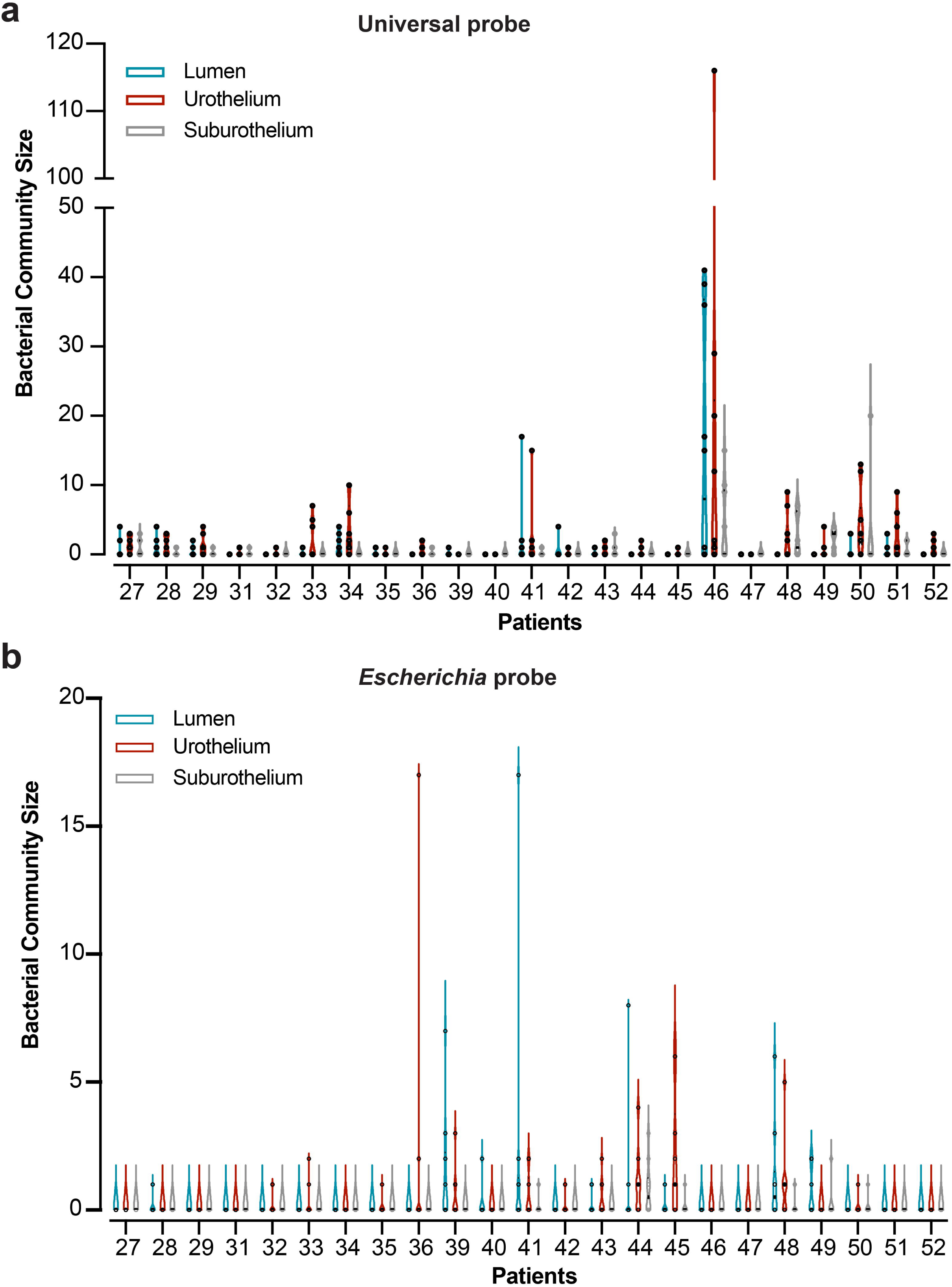
Localization of bacteria detected by Universal and *Escherichia* probes in the bladder wall. Quantification of bacteria detected by Universal (**A**) or the *Escherichia* (**B**) probe in the bladder lumen, urothelium and suburothelium of each patient. Whiskers represent minima and maxima of the data distribution while boxes denote the interquartile range and solid lines denote the median.

Similarly, *Escherichia spp.* were detected by the *Esco 473* probe in 52% of participant biopsies and in 92% of biopsies obtained from patients suspected of having *E. coli* as a causative UTI pathogen from their urine culture history within one year prior to EF (**Table 2**). The largest urothelial and sub-urothelial communities detected by the *Escherichia* probe were observed in PNK036 (N=17) and PNK044 (N=9), respectively (**Figure 4B**). The largest community of luminal *Escherichia* community was observed in PNK041 (N=17).

### Higher bladder bacterial burden is associated with increased risk of UTI relapse

To begin to understand the association between bladder-resident bacteria in rUTI disease progression, we examined the correlation between bladder bacterial burden and rUTI relapse following EF. Since rUTI is defined as two UTIs within six months, we considered participants who experienced a UTI within six months of the EF procedure for this analysis. Eighteen out of 34 (53%) patients had a UTI relapse within six months of the EF procedure (**Table 2**). We observed that patients with higher bacterial burdens had about a 3.1 times greater risk of rUTI relapse within six months (HR = 3.09, 95% CI: 1.16-8.74; *p* = 0.025) compared to women with lower bladder bacterial burdens (**Figure 5A**). We then sought to determine if *Escherichia-* specific bladder burden was associated with a higher risk of rUTI relapse but observed no evidence of an association between higher *Escherichia* bladder burden and increased risk of UTI within 6 months post-EF (HR= 2.14; 95% CI: 0.58-7.90; *p* = 0.26) (**Figure 5B**). An exploratory analysis was performed to determine if any other variables were associated with the risk of rUTI relapse. Age (**Figure S2A**), Diabetes and its types (**Figure S2 C & D)** and stage of cystitis **(Figure S2E)** were not associated with increased risk of relapse, but Universal bacterial burden was significantly associated **(Figure S2B).** We further sought to measure the association between the survival times of rUTI relapse patients and covariates using the Cox proportional hazards model. We found that universal bacteria (i.e., total bacterial burden) was the only variable associated with a higher risk of rUTI relapse (HR: 3.15, 95% CI=1.09 to 9.11, *p* = 0.035) (**Figure S3**). No other variables were associated with both universal bacteria and rUTI relapse, thus all other variables were removed by LASSO.

**Figure 5.**
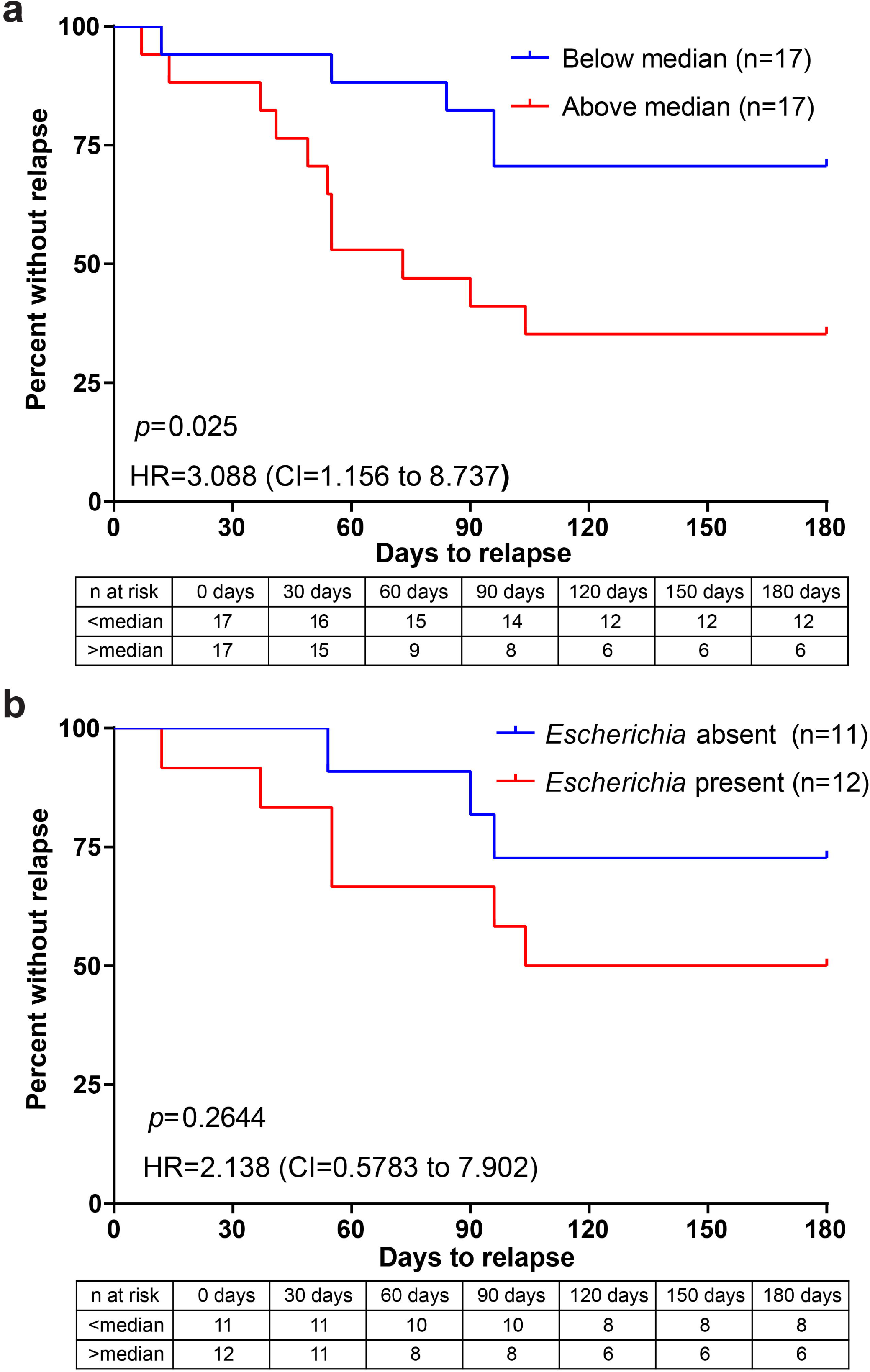
Elevated bladder bacterial burden is associated with a higher risk of rUTI relapse within 6 months of EF. Kaplan-Meier analysis of the association between total bladder bacterial burden as detected by the universal probe (**A**) or *Escherichia* burden (**B)** and risk of UTI within six months of EF. Risk tables under each plot give the number of participants at risk and the cumulative number of events (A had 16 events; B had 9 events). About 53% of the sample (18 of 34 participants) were right-censored due to not having an event during the observation period. Follow up time Median (IQR): 180 (55-180) days post EF. HR: Hazard ratio; CI: confidence interval.

## Discussion

One hypothesized mechanism for recurrent symptomatic bacteriuria is via reactivation of quiescent intracellular reservoirs (QIRs) (27-32). There is robust evidence for the presence of IBCs and QIRs in mouse models of UTI (9, 33, 34). Justice et al. and Anderson et al. used mouse models of urinary tract infection and found that IBCs were formed by UPEC cells that had invaded bladder epithelial cells and were protected from the host immune system. The IBCs were able to persist in the bladder for several weeks and could cause recurrent infections (35, 36). Mysorekar *et al*. demonstrated the presence of UPEC QIRs within the endosomal compartments of murine bladder superficial and underlying transitional epithelial cells (33, 37).

More recent work has extended the evidence for IBCs and QIRs to human specimens. Rosen *et al.* analyzed human urine samples and found that IBCs were also present within exfoliated superficial cells from patients with urinary tract infections caused by UPEC (30). Using human bladder biopsy samples, De Nisco and colleagues directly observed IBCs and QIRs in the bladders of women with uncomplicated rUTI (16). These bacterial reservoirs were found not only within the urothelium, but within the underlying bladder wall layers and were found to be associated with evidence of chronic inflammation. Recently, Wolfe et al. demonstrated the presence of bacteria in bladder biopsy tissues of females with and without Interstitial Cystitis (IC) (19). The current study extends our knowledge of bladder-resident bacteria associated with rUTI by identifying populations of *Escherichia* within bladder tissue and finding that a higher burden of bladder-resident bacteria is correlated with an enhanced risk of rUTI relapse within 6-months following EF.

This study provides definitive evidence that in some women suffering from uncomplicated rUTI *Escherichia spp.* had taken residence below the urothelial surface of the bladder wall. Surprisingly, given *E. coli* is responsible for nearly 75% of uncomplicated UTI, an increased load of tissue-associated of *Escherichia spp.* as detected by 16S rRNA FISH was not significantly associated with an increased risk of a symptomatic UTI episode within six months following EF (1). In fact, the majority of bacteria detected by the universal probe in the bladder biopsies were not found to be hybridized by the *Escherichia* probe. This finding raises the question of the identity of these bacterial species found deep within the bladder urothelium of rUTI patients and their role in UTI recurrence. Interestingly, a study in a different population of women suffering from IC and not from UTI, found evidence for bladder-associated *Staphylococcus and Lactobacillus,* as well as other bacterial genera, using amplicon sequencing techniques (19). However, these genera have yet to be directly visualized within the bladder wall.

Under the proposed mechanism for antibiotic-refractory rUTI, in which tissue-resident bacterial communities develop within protected niches, one would expect the physical destruction of these communities by an EF procedure to reduce the risk of UTI recurrence. In fact, several clinical studies on EF have confirmed a marked reduction in UTI recurrence after EF (17, 18, 38). However, some women have failed EF, while others have been candidates for repeat EF (39). One possible explanation for these failures from our study findings is that when EF is performed in women with a high load of bladder resident bacteria, these tissue-resident bacteria may be present beyond areas of observable cystitis. Thus, EF may be successful in eradicating the infection at the fulgurated sites but falls short of eradicating tissue-resident bacteria located elsewhere in the bladder, thus possibly explaining UTI recurrence in the 6 months following EF. Another explanation is borne out from our data in that following EF, deeper tissue-resident bacteria from other genera are present and are now unmasked and exposed, allowing the start a new cycle of UTI. Interestingly, preliminary evidence from a separate study indicates that the EF procedure may also alter uroplakin IIIa expression on the urothelial surface (40). It is also interesting that stage of cystitis was not associated with an increased risk of rUTI relapse, but this could be due to the relatively small sample size of our current study. More studies need to be conducted to understand the relationship between short and long-term EF efficacy and bladder-resident bacterial burden in women managed by EF for antibiotic-recalcitrant rUTI.

The main limitations of this study are sample size and diverse degrees of chronic cystitis ranging from early stages to very advanced stages of cystitis when the whole bladder surface is involved (stage 4). Although we used all available metadata to assess potential confounders and did not identify any of these variables as confounding, it is possible that our results may be confounded by unmeasured clinical or demographic variables. Another limitation of the study is that the built-in selection bias of hazard ratios only allows for associative and not causal interpretation (41). Additionally, because of ethics concerns with taking bladder biopsies from healthy individuals, our study did not include a group of healthy human comparators without UTI history. Although we specifically focused on a growing yet underrepresented group of patients, i.e postmenopausal women, future work is needed to expand our findings to other age groups. Furthermore, it is a limitation that 16S rRNA probes can only provide genus or family-level identification and few genus-specific few probe sets have been validated in bladder tissue (42). Future studies should develop more probe sets that can identify other uropathogenic genera as well as genera that comprise the urinary microbiota (43, 44). Once these new probe sets are developed, it will be important to determine the association between the genera information of bacteria embedded within the bladder and those identified in urine cultures from future UTI episodes.

Future studies should also include confirmation that these bladder-resident bacteria are in fact intracellular, and not just residing inside the bladder layers. Additionally, the procurement of bladder biopsies has been mostly restricted to situations when chronic lesions observed on cystoscopy are concerning for early stages of cancer. Several studies have indicated that those situations are rare in women (45-48). However, in light of the data presented here that suggest bladder bacterial burden is associated with rUTI relapse following EF, 16S rRNA FISH of bladder biopsies may serve as a prognostic indicator for clinicians performing EF for the advanced management of rUTI. For example, for patients seeking EF for antibiotic-recalcitrant rUTI, assessment of tissue-resident bladder bacterial load may help to predict their likelihood to respond to therapy or inform the type or duration of antibiotics prescribed after the EF procedure. If additional studies confirm the association of bladder bacterial load and different clinical outcomes after EF, then the quantification of bladder-resident bacteria by FISH serve as a valuable tool for patient stratification and management immediately following the EF procedure that could lead to improved clinical outcomes for the millions of women suffering from antibiotic-refractory rUTI (4, 49, 50).

## Data Availability

All data produced in the present study are available upon reasonable request to the authors

## Author Contributions

Study design and administration was performed by QL, PEZ, NJD. Recruitment was performed by PEZ. The FISH analyses were performed by JGG, PRK. Investigation and interpretation of data was performed by JGG, PRK, FK. Visualization of microscopy results was performed by JGG, PRK. Statistical Analyses were performed by KCL, QL. Supervision and Funding acquisition by PEZ, NJD. Final manuscript preparation by PRK, JGG, PEZ and NJD. All authors reviewed and edited the manuscript. All authors have confirmed that they had full access to all the data in the study and accept responsibility to submit for publication.

## Acknowledgements

The authors would like to sincerely thank the women who participated in this study. This work was supported by a grant from National Institutes of Health (1R01DK131267-01) to N.J.D as well as by the Felicia and John Cain Distinguished Chair in Women’s Health to P.E.Z.

