## Supplemental Material for "Bladder-resident bacteria associated with increased risk of recurrence after electrofulguration in women with antibiotic-recalcitrant urinary tract infection"

Gadhvi *et al.* 2024

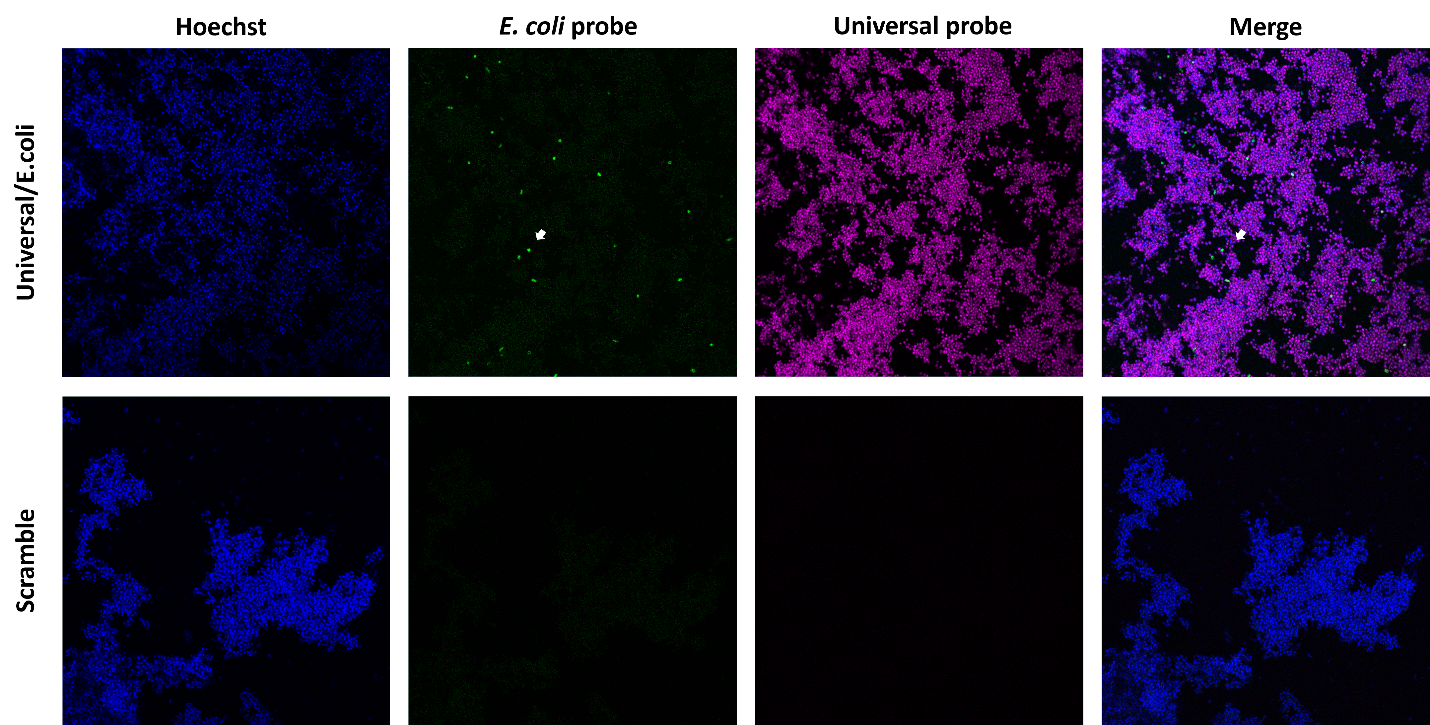

**Figure S1. Validation of *Escherichia* FISH probe on bacteria grown *in vitro.*** FISH was performed on a mixture of *Staphylococcus epidermidis* and *Escherichia coli* in solution that were heat fixed onto the slide. The second panel shows Alexa 488-conjugated *Escherichia* 16S rRNA probe labelling the *E. coli* present in the solution specifically. The panel shows the Alexa 647-conjugated Universal 16S rRNA FISH probe labelling all bacteria on the slide. The bottom panel shows Alexa 488- and Alexa 647-conjugated non-specific scramble probe as negative control.

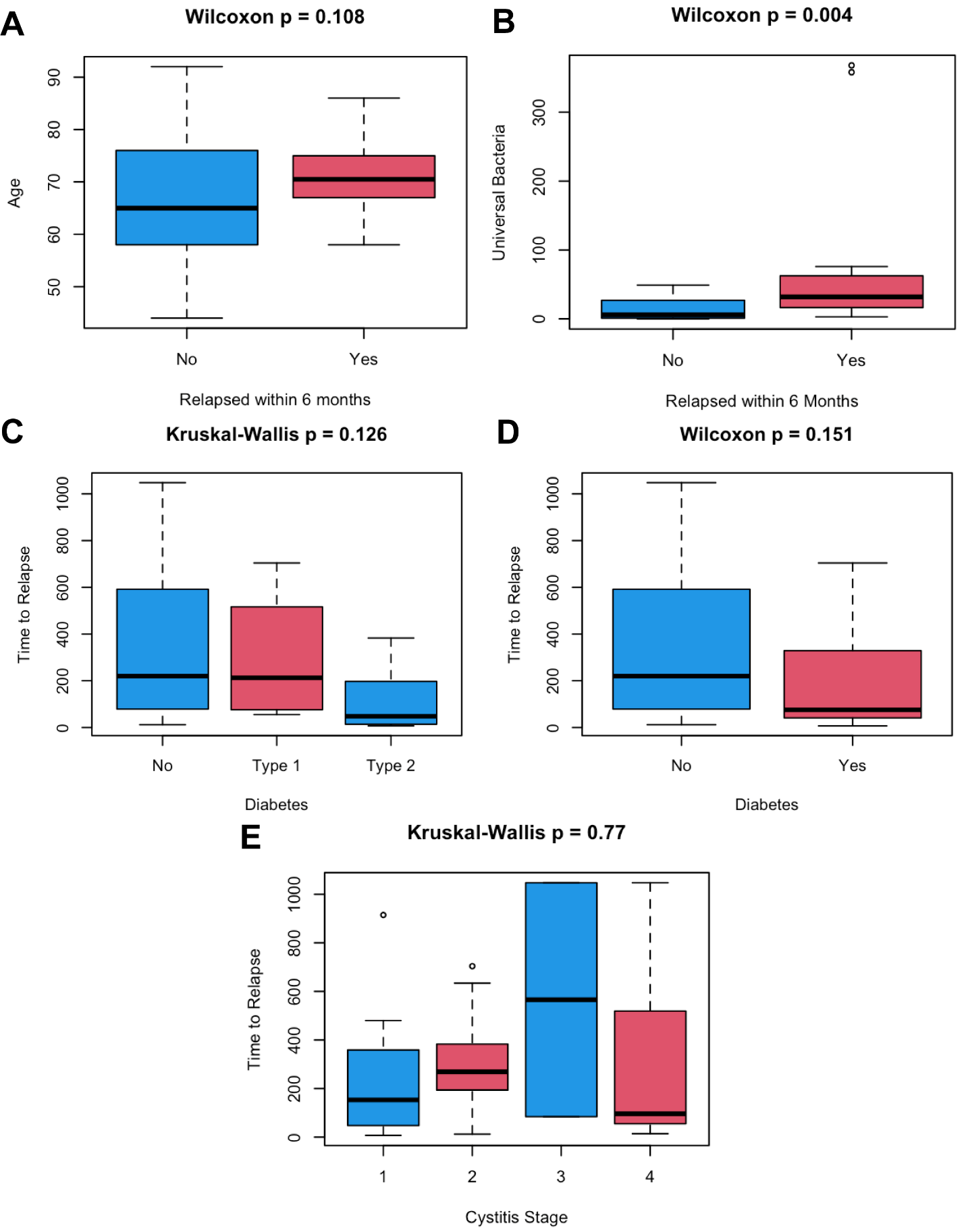

**Figure S2. Exploratory analysis of the association of clinical variables with UTI six months post EF.** Panel (A) Median age is not significantly different between women who experienced UTI within 6 months of EF and those who did not. (B) The number of bacteria detected by the universal probe is significantly elevated in women experience UTI within 6 months of EF. (C) and (D) Diabetes is not associated with time to UTI relapse following EF. (E) Cystitis stage is not associated with time to UTI relapse post EF. Outliers are any data outside of 1.5 times the IQR and are indicated by hollow dots.

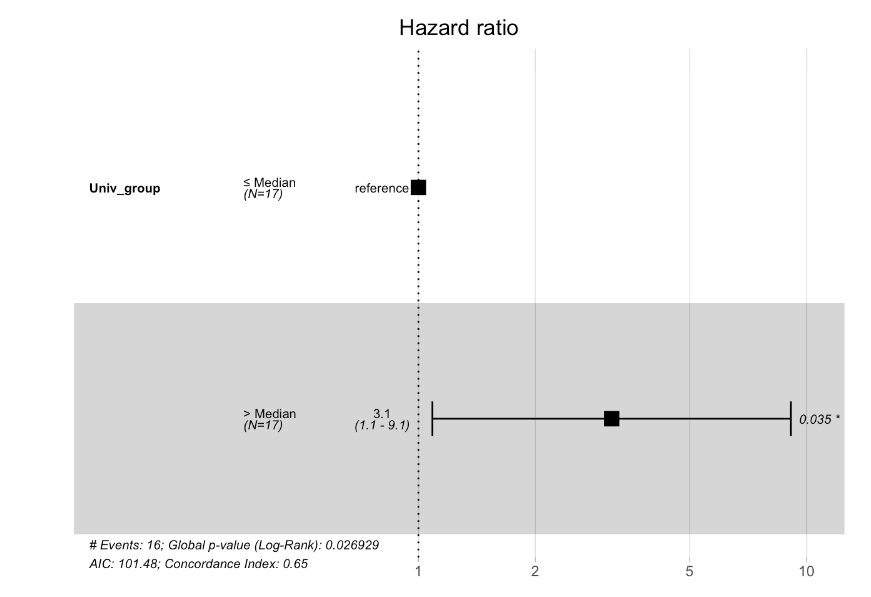

**Figure S3. Cox proportional hazard model shows that universal bacterial burden is the only variable significally associated with UTI relapse withing 6 months.** Compared to other variables like Age, BMI, Diabetes, stages of cystitis, parity and EF history, only the Universal bacterial burden was found to be significantly associated with UTI relapse [Wald test]. The concordance index is 0.65 and the 95% CI for the concordance index is [0.54, 0.76].
